# What association do political interventions, environmental and health variables have with the number of Covid-19 cases and deaths? A linear modeling approach

**DOI:** 10.1101/2020.06.18.20135012

**Authors:** Harald Walach, Stefan Hockertz

## Abstract

**Background and Question:** It is unclear which variables contribute to the variance in corona-virus disease (Covid-19) related deaths and Corono-virus2 (Cov2) cases. We wanted to see which contribution public health variables make in addition to health systems, health, and population variables to explain Covid-19 cases and deaths

**Method:** We modelled the relationship of various predictors (health systems variables, population and population health indicators) together with variables indicating public health measures (school closures, border closures, country lockdown) in 40 European and other countries, using Generalized Linear Models and minimized information criteria to select the best fitting and most parsimonious models.

**Results:** We fitted two models with log-linearly linked variables on gamma-distributed outome variables (CoV2 cases and Covid-19 related deaths, standardized on population). CoV2-cases were best predicted by number of tests (b = 2*10^−7^, p =.00005), life-expectancy in a country (b = 0.19, p < .000001), and border closure (b = −0.93, p = .001). Population standardized deaths were best predicted by time, the virus had been in the country (b = 0.02, p = .02), life expectancy (b = 0.2, p = .000005), smoking (b = −0.08, p = .00001), and school closures (b = 2.54, p = .0001). Model fit statistics and model adequacy were good (model 1: Chi^2^/DF = 0.43; model 2: Chi^2^/DF = 0.88).

**Discussion and Interpretation:** Only few variables were good predictors. Of the public health variables only border closure had the potential of preventing cases and none were predictors for preventing deaths. School closures, likely as a proxy for social distancing in severely ill patients, was associated with increased deaths.

**Conclusion:** The pandemic seems to run its autonomous course and only border closure has the potential to prevent cases. None of them contributes to preventing deaths.

## Introduction

The novel Coronavirus SARS-Cov2 (CoV2) which surfaced in China in December 2019 for the first time created a world-wide pandemic [1, 2] and an associated disease, named Corona-virus-19 (Covid-19), with respiratory stress, heart problems, kidney failures and immunological problems associated with it [3-7]. Countries closed down their borders, their schools, universities and cultural facilities and sometimes even their whole activities. This was due partially to its novelty and its largely unknown properties, but also, because it was soon clear that those infected by the virus could be asymptomatic for up to a week or longer, while still being infectious to others, and because high infectivity, virulence and mortality was assumed.

The spread of the virus was initially very quick following a seemingly exponential growth curve, but abated and the replication numbers went into decline. Currently it is highly debated what contributes to the variance that can be seen both in CoV2 cases, as well as in deaths attributed to Covid-19. While most people assume that political measures have mitigated the spread of the virus [8], others hold that the process is rather autonomous, that the virus recedes after having infected all those in a population susceptible to it and then the infection abates [9, 10]. Moreover, most modeling approaches that were used in early stages of the disease to inform political decision making did not take into account potential inhomogeneity of a population due to natural or specific immunity of a large part of the population [11, 12]. More recent models that take such inhomogeneity parameters into account, informed by novel data, estimated that after about 7 to 18% of a population have been infected herd immunity is reached, because the rest of the population might not be susceptible to the virus [13, 14].

Since it is largely unclear what variables contribute to the variance in cases and deaths attributable to CoV2, we wanted to study this question by building linear models using various predictor variables to study their influence on the outcomes Covid-19 cases and deaths in various countries.

## Method

### Data

We collected data on Covid-19 cases and deaths as presented by the database of the European Center for Disease Prevention and Control on their website on 15th May 2020. We used European and OECD countries, because those data are most relevant to our question and are more validly accessible. We included the following 40 countries

1. Austria
2. Belgium
3. Brazil
4. Bulgaria
5. Canada
6. China
7. Croatia
8. Cyprus
9. Czechia
10. Denmark
11. Estonia
12. Finland
13. France
14. Germany
15. Greece
16. Hungary
17. Iceland
18. India
19. Iran
20. Ireland
21. Italy
22. Japan
23. Lativa
24. Lithuania
25. Luxembourg
26. Malta
27. Netherlands
28. Norway
29. Poland
30. Portugal
31. Romania
32. Russia
33. Slovakia
34. Slovenia
35. Spain
36. Sweden
37. Switzerland
38. Turkey
39. United Kingdom
40. USA

Covid-19 Cases and Deaths were summed for the total period covered by the ECDP-database and used as dependent variables (criterion). We standardized cases and deaths on 100.000 inhabitants, taken from the population size in the same data-base.

As predictors we collated data from publicly available sources (see Supplementary Material for a list and for sources) for population, health, health systems, and environmental indicators between May 15^th^ and 20^th^ 2020. The variables used as predictors are described in a protocol that was published on the server of the Open Science Framework (https://osf.io/x93np/) before commencement of data collection and analysis. Briefly, we used population indicators (Life-expectancy, percent single households, city dwelling, age groups, population density), health systems indicators (number of doctors, hospital beds, ICU beds,

PCR tests), health indicators (percentage of obese persons, diabetes patients, smoking and physically inactive persons), air pollution, and finally variables coding for political actions: closure of borders, closure of schools, country lockdown (all as dummy variables), including the rapidity of implementation since the first case was noted as number of days from first case to date when first action was implemented (see Table 1).

**Table 1.** Nonparameteric correlations of all variables of interest with outcome variables (cases standardized, deaths standardized, and case-fatality rate, defined as number of deaths/cases per 100.000 inhabitants)

|  | Hospital Beds | Doctors | ICU Beds | Number of Tests | Days since first case | life expectancy total | Life expectancy male | Life expectancy female | smoking in total | Smoking male | Smoking female | City-dwelling | Population density | Single household |
| --- | --- | --- | --- | --- | --- | --- | --- | --- | --- | --- | --- | --- | --- | --- |
| Cases | -0,30 | 0,21 | 0,24 | 0,32* | 0,17 | 0,53* | 0,57* | 0,46* | -0,38* | -0,59* | 0,001 | 0,47* | -0,02 | 0,21 |
| Deaths | -0,28 | 0,16 | 0,22 | 0,46* | 0,30 | 0,43* | 0,46* | 0,38* | -0,33* | -0,57* | 0,09 | 0,36* | 0,07 | 0,34 |
| CFR | -0,18 | -0,05 | -0,07 | 0,35* | 0,40* | 0,16 | 0,17 | 0,09 | -0,21 | -0,30 | 0,07 | 0,15 | 0,13 | 0,33 |
| | obesity rates in % | insufficient physical activity | Diabetes | Vaccination | Sleep | Pollution_NO 2 | Pollution BAP | Pollution Ozone | Pollution_PM 2 | Pollution_PM10 | % lipid lowering drugs (\$) | Mercury(\$) | Schools fully closed | Schools partially closed |
| Cases | 0,07 | -0,12 | -0,22 | 0,19 | -0,10 | 0,07 | -0,69* | -0,15 | -0,52* | -0,58* | 0,42 | 0,42 | -0,19 | 0,01 |
| Deaths | 0,04 | -0,07 | -0,31 | 0,12 | 0,06 | 0,31 | -0,49* | -0,11 | -0,39* | -0,42* | 0,42 | 0,40 | 0,10 | 0,08 |
| CFR | -0,01 | 0,05 | -0,34* | -0,02 | 0,13 | 0,39* | -0,13 | -0,02 | -0,01 | -0,05 | 0,41 | 0,19 | 0,31 | 0,19 |
|  | School Closure§ | Duration School Closure | Lockdown# | national lockdown | subnational lockdown | Duration Border Closure | Border closure | Rapidity of political reaction% |  |  |  |  |  |  |
| Cases | -0,07 | -0,06 | 0,01 | 0,07 | -0,07 | -0,10 | -0,43* | 0,11 |  |  |  |  |  |  |
| Deaths | 0,11 | 0,06 | 0,20 | 0,26 | -0,09 | -0,10 | -0,25 | 0,25 |  |  |  |  |  |  |
| CFR | 0,30 | 0,26 | 0,35* | 0,35* | -0,04 | -0,15 | 0,02 | 0,39* |  |  |  |  |  |  |

\$: not usable for modeling because too few data; §: Variable with three categories, taken apart into the dummy variables “Schools fully closed” and “Schools partially closed”; # Dummy variable: 1 if either national or subnational lockdown was present; % Time difference in days between first date a case was registered and first date of a public health intervention
Red/\*: significant; $p < 0.05$

### Statistics

We built two separate linear models to predict the influence of variables on population-standardized CoV2-cases and Covid-19 associated deaths.

In order to investigate which variables might be potential predictors first order correlations of all relevant variables with the outcome variables were calculated, using non-parametric correlations, and their inter-correlation structure was studied. Only variables that contributed with an effect size r > .3 or with significant correlations were further considered for modeling.

As 40 cases offer enough stability to estimate about 4 parameters reliably [15], we opted for small models to start with and included a further predictor only if it was theoretically meaningful, empirically supported (i.e. a significant predictor) and improved model fit. We included first all those relevant predictors from population, health systems, and environmental sets in separate steps that correlate significantly with the outcome and are not collinearly related with each other. We explored model fit to find the best subset for each small group of indicators with forced entry of not more than four variables at a time, retaining only significant predictors for the next step. In a final step we included potential predictors from the set of public health indicators to investigate whether there is any improvement in model fit and whether these variables were significant predictors. The rationale of this procedure is: If the public health measures contribute to preventing cases and deaths, then they would have to emerge as potential significant predictors with negative sign (as they were dummy coded with 1 coding for present and 0 coding for absent). In addition, the model fit of the enlarged model would have to improve.

As an indicator of improved model fit we used the difference of Akaike Information Criterion (AIC in its original and corrected version), the difference of Bayes Information Criterion (BIC) and the Chi^2^-Goodness of Fit test statistic divided by degrees of freedom conjointly to avoid over and underfitting. We always used the model that minimized all of them in combination. To assess model adequacy, plots of predicted versus observed cases, residual distribution plots, and residuals vs. cases were visually analyzed and residual plots were screened for outliers (residuals vs. Chi^2^ statistic).

In a sensitivity analysis the model was recalculated without outliers to see whether the model structure, i.e. the variables used as significant predictors, would be the same and goodness of fit improved. For those sensitivity analyses, AIC and BIC were only used as a further criterion if the difference was large, as the efficiency of these information criteria change with number of cases/degrees of freedom and number of variables [15, 16]. We used Statistica Version 13.1 for all analyses.

## Results

### First order correlations

The nonparametric correlations (Spearman’s Rho) between the two predefined outcome variables, cases and deaths per 100.000 inhabitants, as well as the case-fatality rate (CFR), for illustration, are reported in Table 1.

Of the structural variables describing the health systems only the number of tests conducted correlated significantly with number of standardized cases (r = .32) and deaths (r =.46), as well as with case-fatality rate (r = .35) and with number of ICU beds (r = .39).

Of the variables describing political actions only border closure was negatively and significantly related with standardized cases (r = -.43), but only weakly and non-significantly with number of deaths (r = -.25): Cases tended to be higher in countries that did not close the border. But neither lockdown nor school closures were significantly and sizably related with number of cases or number of deaths. Only full closure of schools was slightly, but non-significantly related with number of cases (r = -.19) but not with number of deaths, indicating that cases were higher in countries that had not closed schools. However, as school closure was correlated positively, but non-significantly (r = .30) with CFR, it is necessary to clarify by modeling, which covariation might be influential.

The duration or length of border- or school closures was only marginally and non-significantly negatively correlated with number of deaths and cases. The rapidity with which countries reacted, i.e. the time difference between the registration of the first case and the initiation of political reactions was only slightly correlated with number of cases, and significantly correlated only with the case-fatality rate (r = .39), i.e. countries that were slower in initiating political actions had a higher case-fatality rate.

Higher and significant correlations were visible with descriptors of populations and health status. There were more cases in countries that had a higher life-expectancy at birth (r =.53), and there were more deaths (r = .43) in such countries as well. There were more cases (r =.47), as well as more deaths (r = .36) in countries with a higher percentage living in cities. However, neither population density of a country, nor percentage living in single households emerged as a potential predictor. Potentially interesting correlations emerged between case-rate and death rate with percentage of population taking lipid lowering drugs (r = .42) and with amount of mercury used in the alkaline-chlorine industry (cases: r = .42, deaths: r = .40). But since we were unable to find enough data for all countries of interest, these variables could not be used for modeling. None of the other variables describing the health status of a population (obesity rate, insufficient physical activity, sleep problems, vaccination rate, percent of diabetes patients in a population) emerged as potential predictors.

Paradoxically, there were more cases (r = -.38), as well as deaths (r = -.33) in countries that had a *lower* percentage of smokers in the population. As this correlation was even higher for male smokers, likely because smoking is predominantly a male phenomenon, the percentage of male smokers was used for further modeling. The same paradoxical relationship can be seen with variables that code for air-pollution, especially with very small particles (PM2 – particulate matter of 2 micron size per m^3^ air), where we see significant negative correlations of r = - .52 with cases and r = -.39 with deaths. Although the correlation with PM10 was somewhat higher, we used PM2 for modeling, because PM2 and PM10 are highly intercorrelated (r = .75) and because we had more cases with data for PM2, most notably USA.

### Modeling

Following our protocol, we constructed a model to account for the covariance structure of the variables. Since the outcome variables, cases and deaths standardized on 100.000 inhabitants per country, showed adequate fit to a gamma-distribution (Figures 1 & 2), we calculated a generalized linear model with a log-link-function on gamma-distributed outcome-variables:

**Figure 1.**
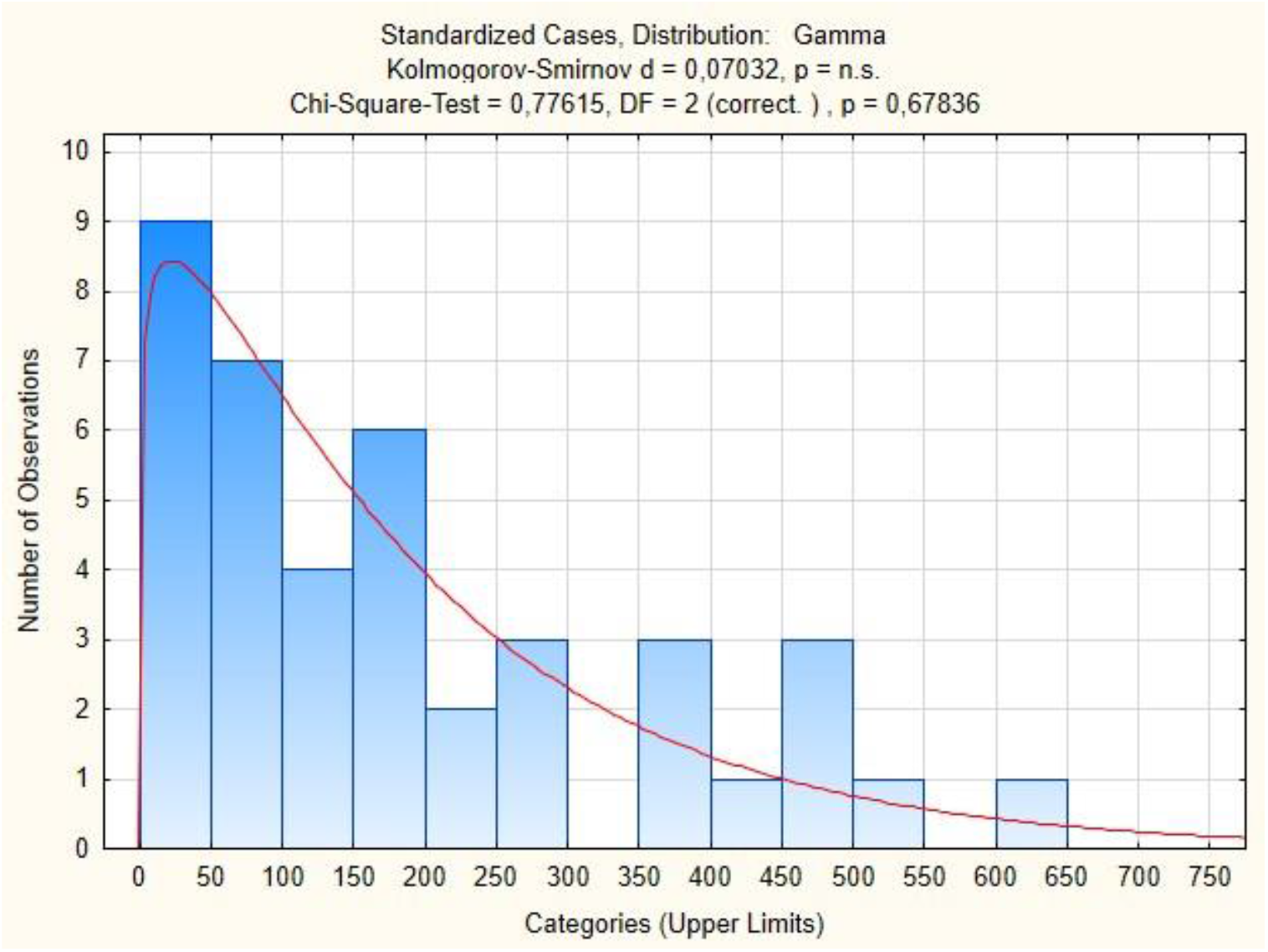
Distribution of CoV2 cases, population-standardized (per 100.000 inhabitants) and approximation to gamma distribution (with Kolmogorov-Smirnov and Chi^2^ Goodness-of-Fit test)

**Figure 2.**
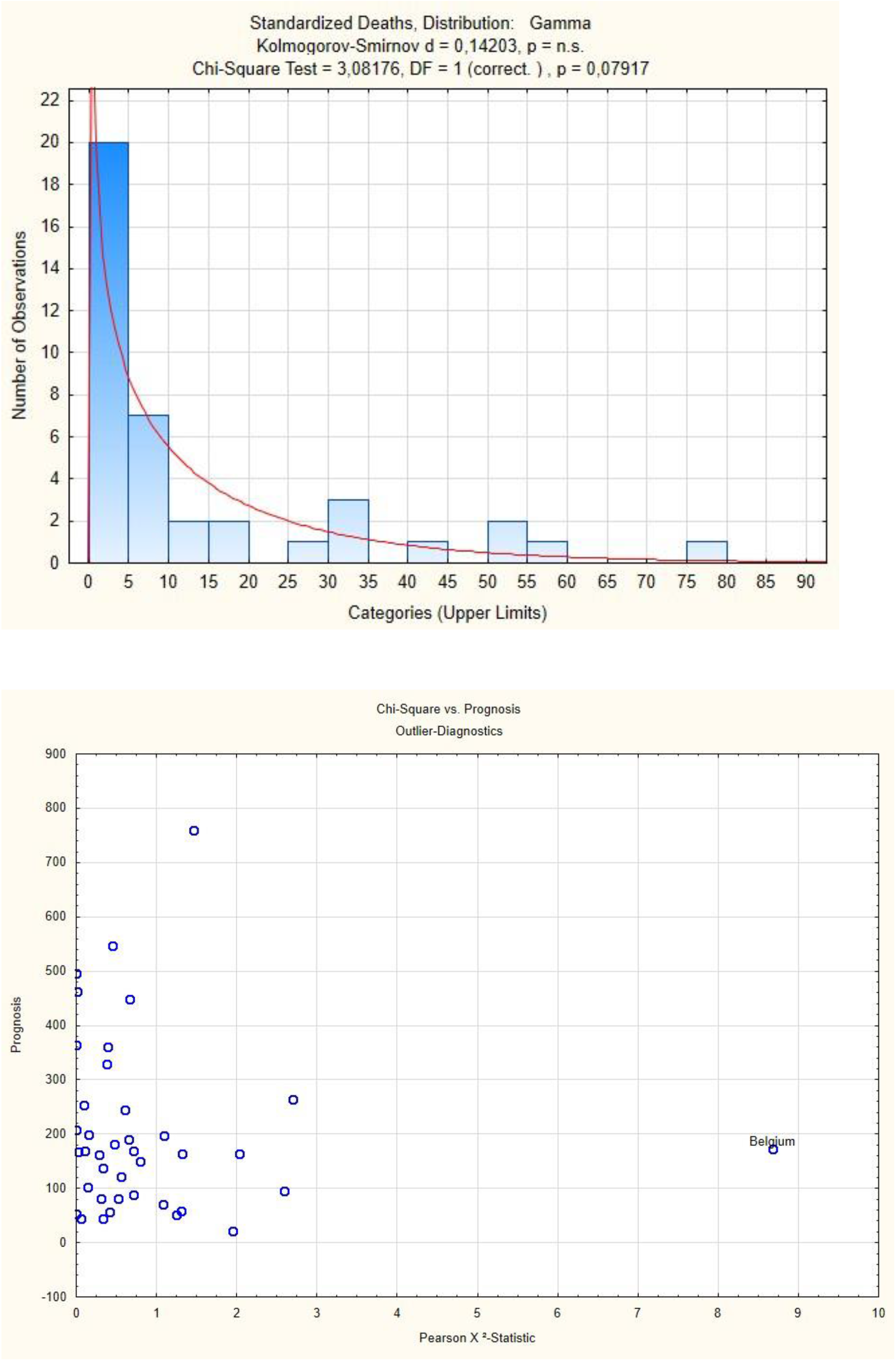
Distribution of Covid-19 related deaths, population-standardized (per 100.000 inhabitants) and approximation to gamma distribution (with Kolmogorov-Smirnov and Chi^2^ Goodness-of-Fit test)

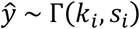

where Γ refers to the Gamma distribution with sh*k*a*i*peand shape*si*, i ∈ 1, …, *𝓃*.

We used the Gamma distribution as it maximizes entropy. Although the (overdispersed) Poisson distribution may have been a choice, we opted for the Gamma distribution because by modeling standardized cases, we are effectively modeling a continuous variable, thereby excluding the Poisson distribution (which models discrete events). We also considered log-transforming the outcome variables to approximate a normal distribution, but the fit was not adequate and sensitivity analyses using linear regression on a log-transformed outcome variable yielded essentially the same results, but with inadequate fit.

### Predicting Cases

The models that best predicted standardized cases are presented in Table 2.

**Table 2.**
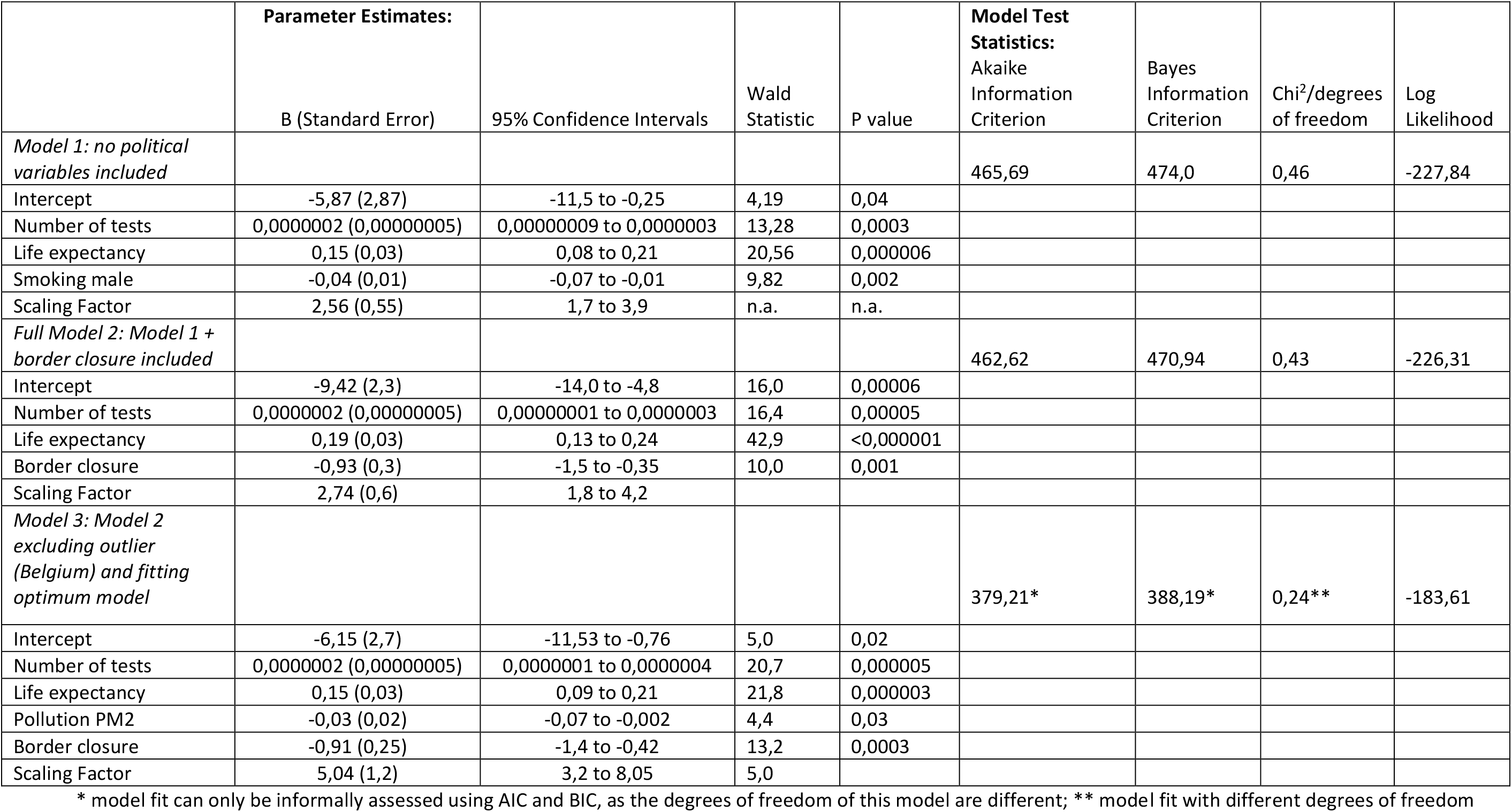
Linear Model Predicting Standardized CoV2-Cases

The first model describes the best fitting model for all countries predicting cases. The variables entering the model are life-expectancy, number of tests and smoking. Parameter estimates are positive for life expectancy and number of tests, and negative for smoking. In a second step variables coding for political decisions (country lockdown, border closure, school closures) were entered. The best fitting model emerged with border closure as a negative predictor, with smoking removed. The model fit statistics show improved model fit over the first model (Akaike Information Criterion – AIC 462,62 vs. 465,69; Bayes Information Criterion 470,94 vs. 474; Chi^2^/degrees of freedom 0,43 vs. 0,46). We inspected the Chi^2^ vs. prognosis plot to spot outliers. There was only one clear outlier, Belgium (Figure 3).

**Figure 3.**
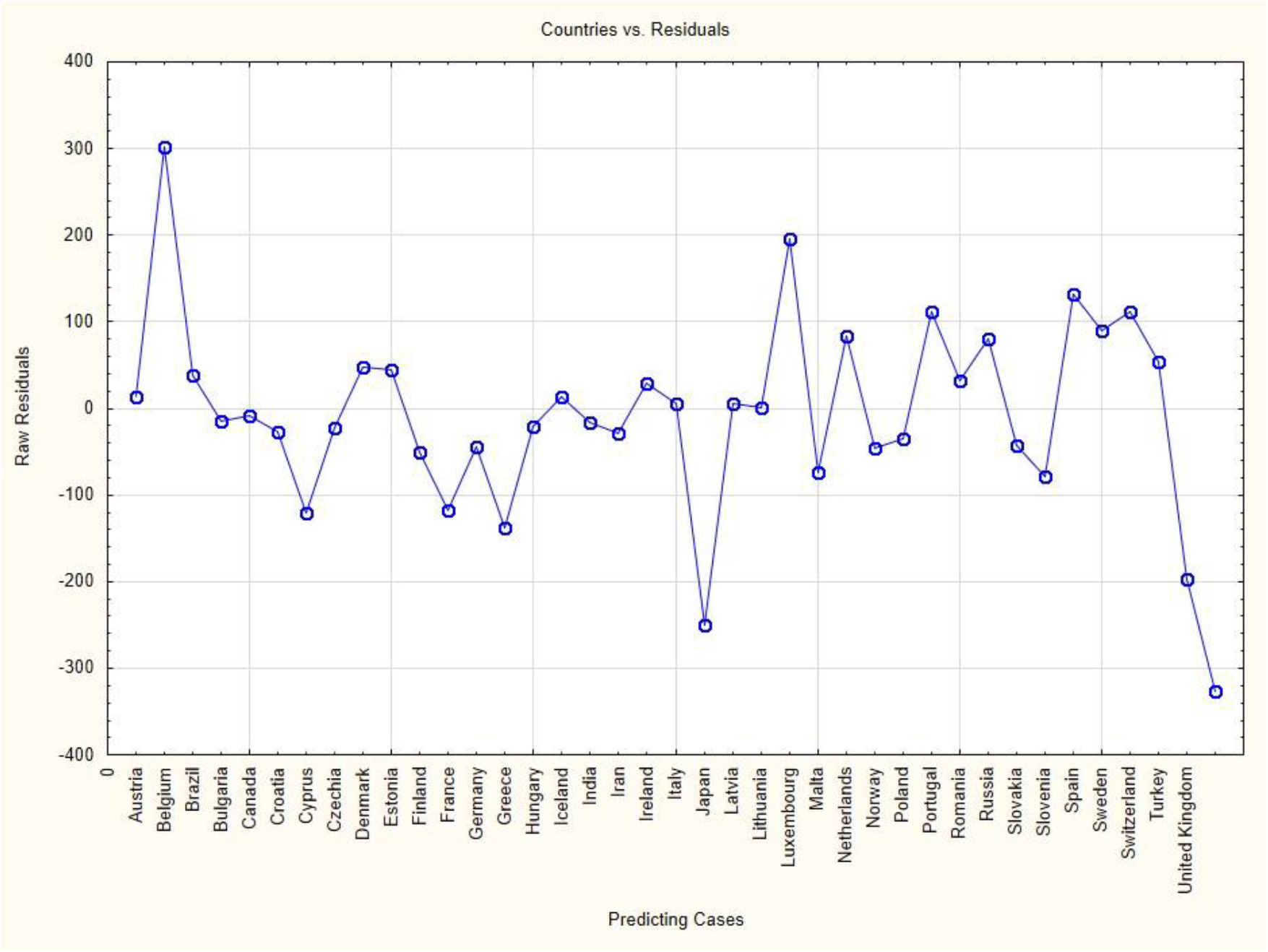
Outlier diagnostic: Chi^2^ vs. prognosis identifies Belgium as an outlier that reduces model fit.

Removing this outlier improved model fit considerably (AIC 379, 21; BIC 388,19; Chi^2^/degrees of freedom 0,24), with air-pollution PM2 added to the model as a negative predictor. The full model can predict the cases in the countries comparatively well. Figure 4 presents the plot of raw residuals against cases/countries for the full model (model 2). In this plot China is missing, because there were no data on PCR-tests for China, and the last country is the USA. The countries for which the model predicts the cases less well, i.e. which have larger residuals are Belgium and Luxembourg with larger positive residuals, i.e. more cases unaccounted for by the model, and Japan, United Kingdom and USA with less cases than accounted for by the model.

**Figure 4.**
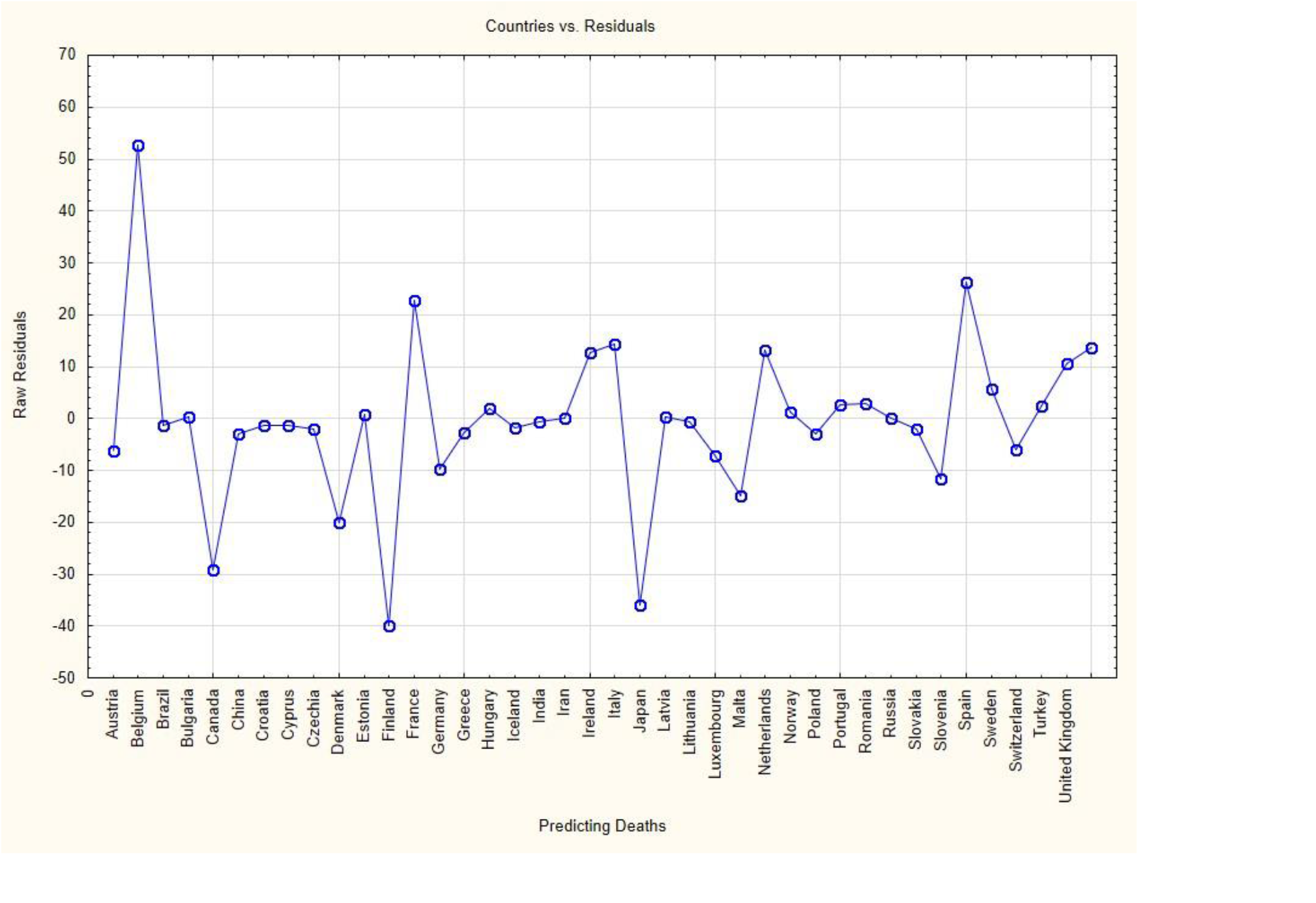
Raw residuals vs. cases/countries for full model predicting cases; the last country is the USA.

### Predicting Deaths

The model predicting Covid-19 related deaths is presented in Table 3: Here the duration the virus had been in the country is a significant positive predictor, and so is life expectancy. Smoking is a negative predictor. When entering the public health variables only school closures emerged as a significant *positive* predictor that improved model fit. Excluding Belgium, the only serious outlier, improved model fit. The same variables remain in the model as significant predictors with nearly the same regression coefficients including their sign.

**Table 3.** Linear Model Predicting Standardized Covid-19 associated Deaths

|  | **Parameter Estimates** |  |  |  | **Model Test Statistics:** |  |  |  |
| --- | --- | --- | --- | --- | --- | --- | --- | --- |
|  | B (Standard Error) | 95% Confidence Intervals | Wald Statistic | P value | Aikaike Information Criterion | Bayes Information Criterion | Chi<sup>2</sup>/degrees of freedom | Log Likelihood |
| *Model 1: no political variables included* |  |  |  |  | 262,32 | 270,76 | 1,02 | -126,16 |
| Intercept | -13,25 (4,12) | -21,3 to -5,2 | 10,3 | 0,001 |  |  |  |  |
| Days of pandemic | 0,02 (0,01) | 0,005 to 0,04 | 7,0 | 0,008 |  |  |  |  |
| Life expectancy | 0,20 (0,5) | 0,1 to 9,29 | 17,1 | 0,00003 |  |  |  |  |
| Smoking male | -0,06 (0,02) | -0,1 to -0,03 | 12,5 | 0,0004 |  |  |  |  |
| Scaling Factor | 1,23 (0,25) | 0,83 to 1,82 |  |  |  |  |  |  |
| *Model 2:*<br>*Model 1 + School closures* |  |  |  |  | 256,47 | 266,07 | 0,88 | -122,24 |
| Intercept | -15,3 (3,8) | -22,8 to -7,7 | 15,8 | 0,00007 |  |  |  |  |
| Days of pandemic | 0,02 (0,007) | 0,003 to 0,03 | 5,3 | 0,02 |  |  |  |  |
| Life expectancy | 0,20 (0,04) | 0,11 to 0,29 | 20,9 | 0,000005 |  |  |  |  |
| Smoking male | -0,08 (0,02) | -0,11 to -0,04 | 19,2 | 0,00001 |  |  |  |  |
| School Closures | 2,54 (0,67) | 1,24 to 3,85 | 14,5 | 0,0001 |  |  |  |  |
| Scaling Factor | 1,4 (0,3) | 1,0 to 2,1 |  |  |  |  |  |  |
| *Model 3:*<br>*Model 2 without outlier (Belgium) and fitting optimum model* |  |  |  |  | 242,97* | 252,94* | 0,84** | -115,48 |
| Intercept | -14,8 (3,7) | -22,1 to -7,4 | 15,5 | 0,00008 |  |  |  |  |
| Days of pandemic | 0,01 (0,007) | 0,0006 to 0,03 | 4,2 | 0,04 |  |  |  |  |
| Life expectancy | 0,2 (0,04) | 0,1 to 0,28 | 20,8 | 0,000005 |  |  |  |  |
| Smoking male | -0,08 (0,02) | -0,1 to -0,04 | 18,1 | 0,00002 |  |  |  |  |
| School Closures | 2,43 (0,65) | 1,1 to 3,7 | 13,8 | 0,0002 |  |  |  |  |
| Scaling Factor | 1,5 (0,31) | 1,0 to 2,2 |  |  |  |  |  |  |

\* model fit can only be informally assessed using AIC and BIC, as the degrees of freedom of this model are different; \*\* model fit with different degrees of freedom

Inspection of residuals show that the linearity assumption is warranted. The model can predict the associated deaths reasonably well (Figure 5). For Belgium, Finland, Japan, and to some extent France and Spain predictions are not so good and the residuals large. Japan and Finland have much fewer deaths that the model cannot account for and Belgium, France and Spain have more.

**Figure 5.**
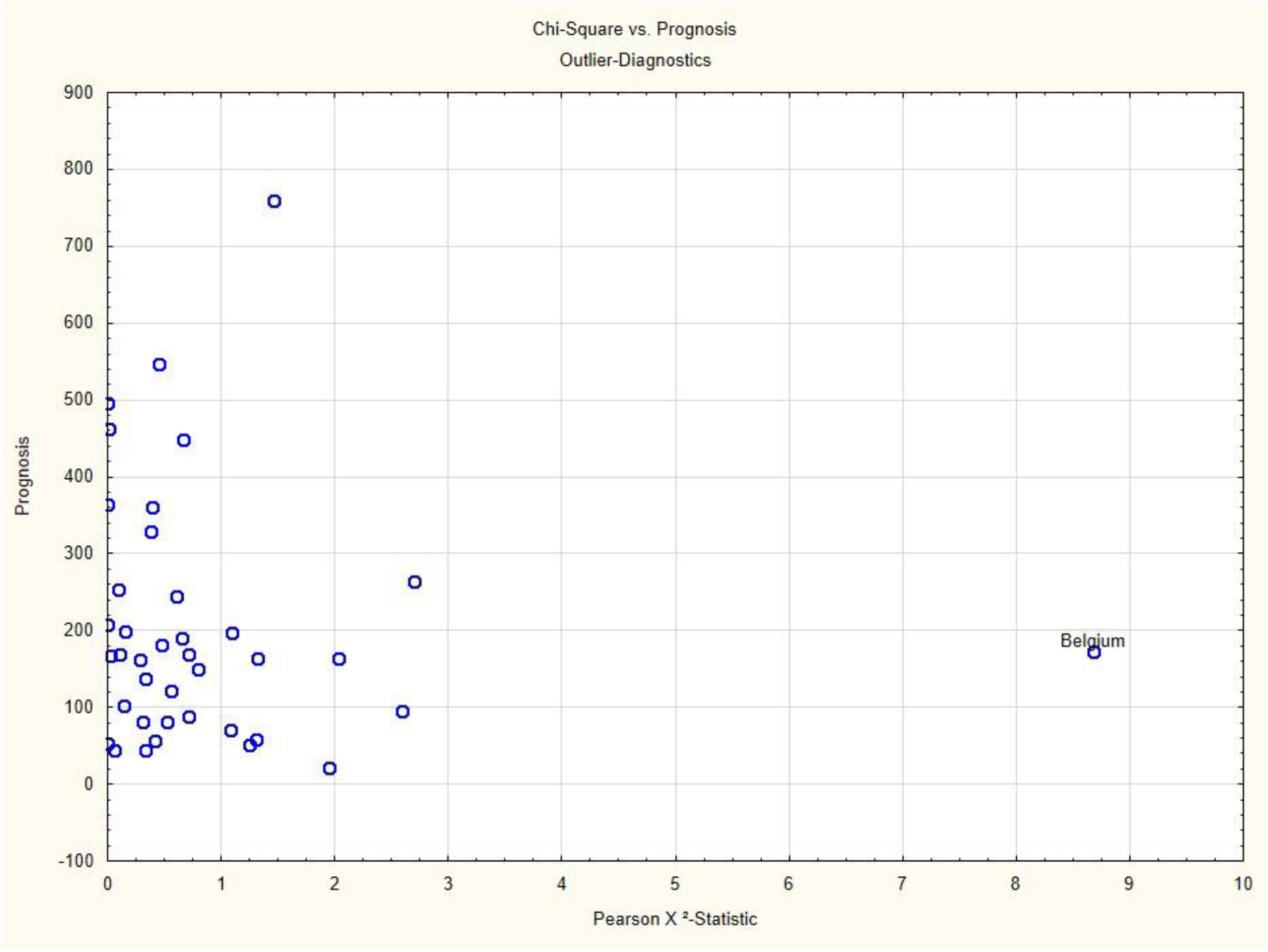
Raw residuals vs. cases/countries for full model predicting Covid-19 related deaths; the last country is the USA.

The inspection of the distribution of the residuals (Supplementary Figure 1) and other diagnostic plots (not shown) confirm that the model assumptions are not violated and the linearity assumption holds.

## Discussion

The major findings of this modeling study using population data for 40 countries are clear: Life-expectancy emerges as a stable positive predictor both for standardized cases of CoV2 infections, as well as for Covid-19 related deaths. Surprisingly, smoking emerges as a stable negative predictor, i.e. protective factor. Of the public health or political variables only border closure is as a strong negative predictor for cases. But school closures are a strong positive predictor for deaths, i.e. is associated with more deaths. The parameter for number of tests conducted in a country is as a strongly significant positive predictor.

The fact that life expectancy is the most consistent positive predictor – the longer the life expectancy in a country the more cases and deaths – is easy to understand. The disease affects most aggressively elderly and multimorbid persons. Life expectancy is a complex variable, incorporating social and medical progress in a country as well as economic indicators, and hence it denotes the number of the elderly in a population, as well as the intensity of medical care. Only the number of CoV2-PCR-tests, out of all health systems variables, enters the model as a significant positive predictor. Other quality indicators of the medical system (number of doctors per 10.000 inhabitants, number of ICU or hospital beds) do not enter the model. That number of tests should be related to the number of cases is evident: the more tests are conducted in a country the more cases can be potentially registered.

The absence of other indicators from the set of medical system variables shows that the development both of infections and deaths is rather independent of the preparedness of the medical system. Although some interesting first order correlations indicated that health status variables might be interesting to explore, none of them came up as a predictor, except smoking as a somewhat protective variable. This might have to do with the fact that smokers have a hyperactive system to combat airborne noxes and hence might have a small advantage against this particular disease [17, 18]. A large cohort study has documented a similar counterintuitive effect [19], and an argument could be made that this might have to do with the fact that smokers express fewer ACE2 receptors [20], which are the main entry gate of CoV2 into the lungs. [2] However, the correlation of smoking with life expectancy is negative (r = -.50 for men), and hence smoking might confer other risks that shorten lives. Only in one model, excluding the outlier Belgium, does air pollution play a role as a potential negative predictor, i.e. as a potential preventive factor. This is rather counterintuitive. Either it could be understood along the same lines: lungs prepared to deal with small noxes might be better prepared to fight a virus. Or else, airborne viruses might be captured by small airborne particles and might fall to the ground earlier. Again, this could be an accidental effect that should not encourage air pollution, as this has detrimental effects elsewhere.

The closing of borders is a significant negative predictor, denoting a protective effect, in the model including all countries and predicting number of cases, but not for predicting number of deaths.

In all models predicting death the time the disease had been in a country is a positive predictor. As deaths develop with a delay of perhaps 3-4 weeks after the first contact with the virus [21-24], this relationship reflects a quite independent temporal dynamic of the infection. It is interesting to observe that closure of schools emerges as a strong positive predictor for the number of deaths, i.e. school closures are associated with *more* deaths. This could be an indicator for strong social distancing rules in a country which might be counterproductive in preventing deaths, as social distance for very ill, and presumably also very old patients, might enhance anxiety and stress and could then become a nocebo [25, 26]. It could also reflect the fact that countries which saw a rising tendency of deaths closed schools as an emergency measure, and hence school closure is an indicator of fear in a country. But considering the prevention of deaths, none of the public health measures studied are associated with the prevention of deaths. Border closure might be an exception in that it is associated with reduction in the number of cases and hence, indirectly, the number of deaths. But in the full model predicting deaths it is not a significant predictor. This seems to contradict new modeling data using time series models [27, 28] that report clear evidence for the effectiveness of non-pharmaceutical interventions. We doubt the validity of these findings. The major shortfall of these models is that they ignore the most likely reason why we find the data we find: immunity in the population and neglecting the strength of natural immunity (see below). Apart from this these models operate with infection-fatality rates (IFR) of 0.91-1.26, a figure derived from early estimates, which is clearly too high. Meanwhile, better estimates are available, and a recent review of seroprevalence studies estimated a mean IFR of 0.25% across 23 studies and all strata of age, and 0.04% for those younger than 70. [29] Along the same line, a new reliability study of such models shows that they are crucially dependent on parameters assumed and the time point at which they capture data [30]. If the wrong assumption about a potential resistance against an infection in a population is made, the results are far off from true values.

Rapidity of reaction can be a positive predictor in some models, but reliably leaves the equation, as soon as the duration of infection is taken into account. This signals in our view, the fact that the dynamics of the infection develops quite independently of political actions, or rather that political actions are mostly too late.

The time the disease had been in a country was only a significant predictor for standardized deaths. A model including this variable to predict standardized cases is not significant and does not improve model fit.

One might argue that a perhaps more conventional way of modeling would have been to log-transform the outcome variables and use standard linear regression approaches. We tried this as a sensitivity analysis but did see essentially similar results with a residual distribution that signaled model inadequacy, and hence we doubt that such a model would have helped with understanding the data better.

As data on number of tests were not available for China one might argue that our model is inadequate, as it excludes an important country. However, we fitted models without the number of tests as a predictor which did not lead to better fit or more meaningful models. The fact that the model predicting deaths, where China is included, could predict cases in China nearly perfectly shows indirectly that this is not an issue. We also used population standardized tests as an alternative to raw number of tests, but found that the model fit was much worse.

Thus, the image that emerges from the data and the attempt to understand their relationship through modeling is that of a largely autonomous development. It affects mainly the elderly. Smoking is somewhat protective and border closures is associated with a lower number of cases. But other measures – closing of schools and lockdown of whole countries – do not contribute to a reduced number of cases or deaths. This may have to do with the fact that the virus travels extremely quickly. Even the shutdown of Wuhan airport delayed the spread across China only by 2,8 days [31, 32], and as Chinese airports remained open the spread of the virus across the world was guaranteed and could not be stopped by gross measures such as border closures, as these came too late. An Italian seroprevalence study estimated that even at the very beginning of the pandemic in Italy, the first country to be affected in Europe, there were 2.7% of the population in Milan that had already had contact with the virus. [33]

The examples of Taiwan [34] and Hongkong [35] show that containment is possible, if reactions come very quickly and if cases can be traced close to 100%. But already the presence of 5 cases in a population increases the likelihood of a pandemic by 50%. [36, 37] Once infections are in the vulnerable segments of a populations, like in hospitals or homes for the elderly, political actions like school closures or country lockdowns do not prevent deaths. What might be useful but cannot be seen in our coarse-grained data are special protective measures geared to protect these vulnerable populations, such as protective masks for personnel and visitors in hospitals and old people’s homes, or the wearing of face masks in places with bad ventilation and close proximity of people.

Why, then, have infections subsided and deaths receded since we gathered our data on May 15^th^ 2020? Most people would say this was due to the public health measures [1], and recent modeling studies seem to support this [8, 27, 28]. However, we have pointed out that the peak of the cases had been reached in Wuhan already on January 26^th^, only 3 days after the city lockdown. [38] This was surely too short to be a result of public health measures, as cases manifest with a delay of at least 5, rather more days. And a careful analysis shows that, if one uses realistic retrodiction of cases, then effects of public health measures cannot be seen [39]. Thus, our modeling supports the view that the public health measures of school closures and country lockdown, with the exception of the closure of borders to reduce cases, were likely ineffective in influencing cases and deaths. If anything, social distancing might even be harmful for seriously ill patients.

Very likely, scientists and governments overestimated the danger this virus presented and underestimated the immunological resistance in the population. While there is no doubt that those really falling seriously ill from this infection suffered a lot more and were in much greater danger than comparable patients suffering from flu or other respiratory infections [22], there can also be little doubt that basic immunological insights were neglected from the outset. Both specific [29, 40, 41] and non-specific immunity [42-44] seems to have been much greater in the population than initially assumed. This is likely the case because the difference of CoV2 from other Corona-viruses is not as great as initially assumed. Thus, a considerable percentage of any population would have been immune through specific cross-immunity against other corona-viruses, apart from the fact that non-specific immunity has been neglected in the discussion nearly completely. This is the reason why more recent models that account for this fact and introduce inhomogeneity parameters reach the conclusion that it is sufficient if 7%-18% of a population have had contact with CoV2 to reach herd immunity [13], and that further waves are unlikely given immunity [45].

Our data are not foolproof but first important hints. We were unable to code more countries, due to restrictions in time, resources and availability of data. This reduces the stability of estimates and to some degree also variance, although for those variables of interest variance was large enough to estimate stable models. [46] For the chosen models the goodness of fit test signals good fit, and the relative improvement of AIC and BIC values from model 1 to model 2 is obvious. [47-49]

We opted for completeness of data as much as possible rather than for a large number of countries, as modeling depends on the completeness of case-wise data. Some interesting and potentially useful predictors we were unable to gather: a more fine-grained resolution of different social distancing rules in different countries, availability and wearing of face masks for medical personnel and the public, for instance. Every model is wrong [50], but given the data our models have a comparatively good fit. This can also be seen indirectly, as excluding an outlier improved model fit, but did not change the predictors and their overall structure. Also, the plot of residuals versus cases/countries shows that the models fit most countries well. In Belgium, Japan, Luxembourg, United Kingdom and USA other factors seem to play a role that were not fully captured by these predictors, when predicting cases. In predicting deaths again Japan, but also Finland are not well predicted with fewer deaths, and Belgium, Spain and France seem to have more deaths which our model cannot account for.

Obviously, in a population-based study we have to rely on the validity of the data provided by other sources, which may be of variable, even doubtful quality. This limitation has to be borne in mind. We gleaned our data from respectable sources, and inspecting our residuals vs. countries plots (Fig. 4 and 5) we can see that large countries with potential testing and reporting problems (Brazil, Russia, China, Iran) are nearly perfectly accounted for by our model, and hence unreliability of data does not seem to be a major problem.

An exploratory modeling approach is always open to critique, as it is an observational study trying to infer potentially causal factors from a cross-sectional piece of data. This has to be borne in mind. We decided to build models that are theoretically guided and conceptually informed [15], starting with health systems, structural and population indicator variables and entering political public health variables in a last step, then adapt the model to find the best model fit. We followed a predefined, published protocol which guarded us against aimless fishing, and strove for parsimonious models that could explain the data with a minimum of predictors and good model fit. We avoided computer guided step-down and step-up procedures as they are inefficient or prone to overfitting. [15] Thus, we are quite confident that we did not overlook an important contribution of political actions to an explanatory model: they are not visible in our data except for those we report. More fine-grained, or country specific analyses might eventually unravel some contribution of such procedures, but on a large scale population based level they are not visible apart from those we report.

## Conclusion

In our data-set of 40 countries, only border closure had the potential to prevent cases. Other public health measures were not associated with reduced CoV2-cases or Covid-19 associated deaths. Rather, the pandemic seems to take its own course. Since being elderly is a risk factor that cannot be changed, for many diseases and death, political actions in future pandemics would likely need to focus on protecting these members of society first. Apparently, closing schools and locking down countries was not the right method to prevent deaths. Perhaps the most sensible measures against pandemics are high alertness and an early warning system that initiates rapid actions that can prevent pandemics from developing.

## Data Availability

The data will be made publicly available on the Open Science Framework upon publication and to peer reviewers and other qualified persons beforehand.

## Acknowledgement

We are grateful to Sebastian Sauer for advice on modeling and for counter-checking the validity of our modeling with R-routines and for critically commenting on an earlier draft. We thank the students of the master class “Quantitative Research Methods” of the MSc course “Health Promotion” of the University of Applied Sciences in Coburg, Germany, who gathered the data for this study and participated in discussing and initiating this project. We are grateful to Karin Meissner for advice and some references.

## Declarations

### Ethics

Not applicable for secondary data-analysis.

## Appendix: Supplementary Material

**Figure S1.**
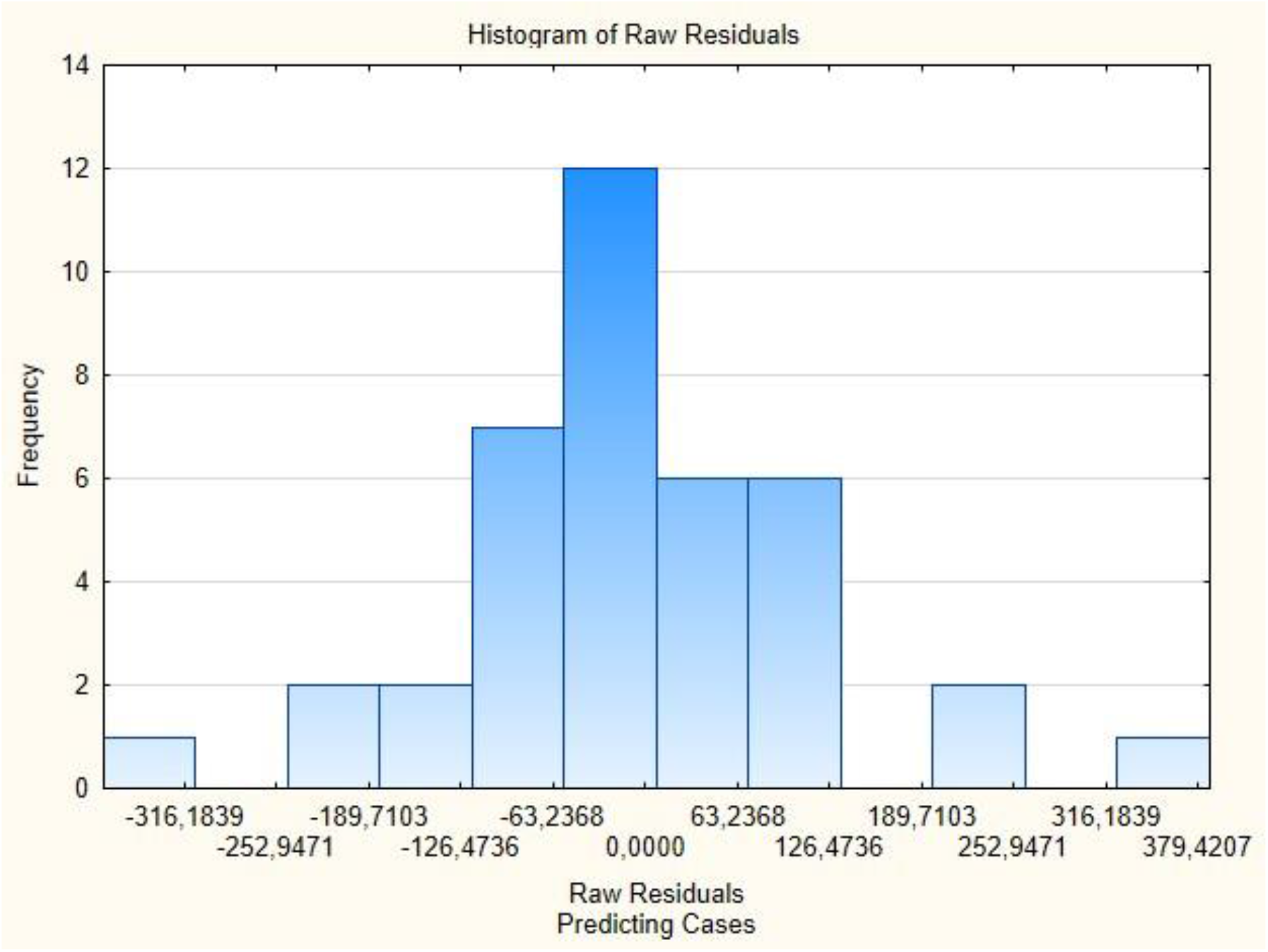
Model diagnostic: histogram of raw residuals - predicting cases.

## Supplementary Material S2: Data-Sources, Data-Treatment and Analysis Log

### Data Extraction: Inclusion and Exclusion

Data were extracted from the the Database of the European Center for Disease Prevention and Control, as of 15^th^ May 2020. This yields day-wise cases, deaths and population numbers for each country. The data were parsed (cases and deaths summed, date of first case registration and populations numbers extracted) using the Statistica Data Reporting Tool and the countries of interest included. These were all European countries including Switzerland, as well as other countries of the OECD, including China, Iran, Russia, India, Japan and Brazil to represent all countries that were at the beginning of the crisis, as well as other large countries in the world. We excluded Africa, other South and Middle American and Asian countries because of a lack of resources, time and because we were not sure we would be able to find sufficient data. This yielded the 40 countries described in our protocol.

#### Political Data

**Data on border closure** came from the European commission:

European Commission. Mobility and Transport. Coronavirus Response. Access on18.05.2020: https://ec.europa.eu/transport/coronavirus-response_en

For the following countries we used other sources, as these were not contained in the European Commission data base:

*Brazil, Canada, Japan, Lithuania, Switzerland, Turkey, USA* : https://www.nytimes.com/article/coronavirus-travel-restrictions.html

*India*: https://www.cnbc.com/2020/03/12/coronavirus-india-suspends-most-visas-closes-land-border-with-myanmar.html; all accessed on 18.05.2020

We checked political data against Wikipedia (country lockdowns, border closures): https://de.wikipedia.org/wiki/COVID-19-Pandemie#/media/Datei:COVID-19_Outbreak_lockdowns.svg

Data for border closure of Iran suggested that other countries closed their borders against Iran, but not Iran against other countries. Hence this was marked as open.

We wanted to include the availability of face masks and their number, but those data were unavailable for most countries, so we excluded this variable.

### School Closures

Were taken from the UNESCO data base which updated them on a daily basis; we extracted the data on the 15^th^ of May:

UNESCO. COVID-19 Educational Disruption and Response. Access on 18.05.2020. https://en.unesco.org/covid19/educationresponse

### Health and Environmental Data

We extracted data for ***vaccination rates*** from https://de.statista.com/statistik/daten/studie/1034782/umfrage/laender-mit-d,er-hoechsten-impfquote/

<u>This is a representative survey and covers all different vaccinations. As this covers only Europe, we searched other sources. For China, India, Iran, Japan data came from</u> https://ourworldindata.org/grapher/immunization-coverage-against-diphtheria-tetanus-and-pertussis-dtp3-vs-gdp-per-capita

For Brazil from https://academic.oup.com/jtm/article/25/1/tay100/5127106

For Canada from https://www.canada.ca/en/services/health/publications/vaccines-immunization/vaccine-uptake-canadian-children-preliminary-results-2017-childhood-national-immunization-coverage-survey.html

#### Air pollution

Data for air pollution are available for Europe from: https://www.eea.europa.eu/de/themes/air/intro

Air pollution for other countries:

Brazil (2018) : https://www.scielo.br/scielo.php?pid=S0103-50532020000300523&script=sci_arttext 2018 No2, o3, p10, p2.5

Canada (2018): https://pollution-waste.canada.ca/air-emission-inventory/

China, only PM2.5 from 2015 http://berkeleyearth.org/wp-content/uploads/2015/08/China-Air-Quality-Paper-July-2015.pdf4

<u>USA from</u> https://www.epa.gov/outdoor-air-quality-data/air-quality-statistics-report

<u>CSV formatted for all counties for 2017; imported into new spreadsheet and calculated median across PM2.5 weighted 24 h average, because there were a few counties with very high values, median was taken, else it would have been 103 instead of 40. Only PM2 and PM10, other values are not compatible (no full daily or annual averages).</u>

<u>Data including US air-pollution data Covid10-mastertabelle10.sta</u>

<u>Some countries missing on that variable.</u>

### Life expectancy since birth

https://ec.europa.eu/eurostat/databrowser/view/tps00205/default/table?lang=de (18.05.2020)

Brazil, Canada, China, India, Iran, Japan, Russia and USA https://www.laenderdaten.info/lebenserwartung.php#by-population (18.05.2020)

### Smoking status

https://www.who.int/data/gho/data/indicators/indicator-details/GHO/age-standardized-prevalence-of-current-tobacco-smoking-among-persons-aged-15-years-and-older (18.05.2020)

### Physical activity

https://www.thelancet.com/journals/langlo/article/PIIS2214-109X(18)30357-7/fulltext Regina Guthold, Gretchen A Stevens, Leanne M Riley, Fiona C Bull, Worldwide trends in insufficient physical activity from 2001 to 2016: a pooled analysis of 358 population-based surveys with 1·9 million participants, The Lancet Global Health, Volume 6, Issue 10, 2018, Pages e1077-e1086, ISSN 2214-109X, https://doi.org/10.1016/S2214-109X(18)30357-7.

### Obesity rate

https://www.who.int/gho/publications/world_health_statistics/2020/EN_WHS_2020_TOC.pdf

World health statistics 2020: monitoring health for the SDGs, sustainable development goals. Geneva: World Health Organization; 2020. Licence: CC BY-NC-SA 3.0 IGO.

## Sleep Problems

Were exctracted information from various reviews and publications:

Self-assessed “Problems Sleeping” from a review on Statista (https://de.statista.com/statistik/daten/studie/888175/umfrage/selbsteinschaetzung-zu-ausreichend-schlaf-in-ausgewaehlten-laendern/) and https://www.who.int/news-room/detail/14-05-2020-substantial-investment-needed-to-avert-mental-health-crisis

Prevalence of insomnia from various review sources:

Canada: Morin et al. (2011); Portugal: Ohayon & Paiva (2005); China: Xiang et al. (2008); Switzerland: Angst et al. (1989); Greek: Paparrigopoulos et al (2010); Ireland: Nugent et al. (2008); UK: Morphy et al. (2007); Japan: Kim et al. (2000); India: Panda et al. (2012); Brazil: Lopes et al (2014); Spain: Ohayon & Sagales (2010)

Incidende of insomnia in Europe:

Riemann, D., Baglioni, C., Bassetti, C., Bjorvatn, B., Dolenc Groselj, L., Ellis, J. G., … Spiegelhalder, K. (2017). European guideline for the diagnosis and treatment of insomnia. *Journal of Sleep Research, 26*(6), 675-700. doi:10.1111/jsr.12594

Sleep dissatisfaction from Canada: Morin et al. (2011); Finland, France, Turkey, Latvia, Lithuania, Malta, Greece, Ireland, Norway, Spain: May et al. (2018); Portugal: Phayon & Paiva (2005); Switzerland: Statista “Leiden Sie unter Schlafstörungen?”(2019); USA, France, Germany, Italy, UK, Japan: Léher et al. (2007).

And prevalence of sleep problems from Van de Straat & Bracke (2015); Iran: Hosseini et al. (2018); USA: Olufunmilola et al. (2017); Norway: Pallesen et al. (2014); Japan, India, China: Gulia & Kumar (2018).

As these sources gave different types of data for incomplete sets of countries, but together most countries were covered, we used the following strategy:

New variables were constructed, in which the respective original variables were rank-ordered. Then a new hyper-variable was constructed in which the mean rank of those variables that were available per country was deposited. Finally, this new mean rank-variable was again ranked to yield the rank order of countries with sleep problems. This was used for further analysis.

## Health Services Data

We exctracted number of doctors, standardized per 1.000 inhabitants, number of hospital beds and number of ICU beds, standardized.

## Hospital Beds (per 1.000 inhabitants)

WHO: Global Health Observatory data repository. Hospital bed density. Data by Country. Last updated 2020-03-10. Zugriff am 15.05.2020. Online verfügbar unter https://apps.who.int/gho/data/view.Mayn.HS07v

## number of doctors (per 10.000 inhabitants)

Global Health Workforce Statistics. Medical doctors. World Health Organization, Geneva. Last updated 2020-02-14. Zugriff am 15.05.2020. Online verfügbar unter https://apps.who.int/gho/data/node.Mayn.HWFGRP_0020?lang=en

## According to the year given in the data base

Global Health Workforce Statistics. Medical doctors. World Health Organization, Geneva. Last updatet 2020-02-14. Zugriff am 15.05.2020. Online verfügbar unter https://apps.who.int/gho/data/node.Mayn.HWFGRP_0020?lang=en

## Number of Intensive Care Unit Beds

Taken from the OECD report “Beyond Containment: Health systems responses to COVID-19 in the OECD” (p.13 https://oecd.dam-broadcast.com/pm_7379_119_119689-ud5comtf84.pdf); accessed 15^th^ May 2020; see also: http://www.oecd.org/coronavirus/policy-responses/beyond-containment-health-systems-responses-to-covid-19-in-the-oecd-6ab740c0/

The following countries are not contained in this list and data for these countries come from the according sources:

China: *Phua, J*.; *Farug, M*.; *Kulkarni, Atul; Redjeki, Ike* (*2020-01-01*). *“Critical Care Bed Capacity in Asian Countries and Regions”. Critical Care Medicine. doi:10.1097/CCM.0000000000004222*.

Czech Republic, Estonia, Finland, Greece, Iceland, Latvia, Lithuania, Luxembourg, Portugal, Slovakia, Slovenia, Sweden: *Rhodes, A*.; *Ferdinande, P*.; *Flaatten, H*.; *Guidet, B*.; *Metnitz, P. G*.; *Moreno, R. P*. (*2012-10-01*). *“The variability of critical care bed numbers in Europe”. Intensive Care Medicine. 38* (*10*): *1647–1653. doi:10.1007/s00134-012-2627-8. ISSN 1432-1238. PMID 22777516*.

Germany: *DIVI-Intensivregister https://www.intensivregister.de/#/intensivregister*

Russia: *http://government.ru/news/39218*

Turkey: *https://dosyamerkez.saglik.gov.tr/Eklenti/33116,haber-bulteni2018-30092019pdf.pd*

## Number of Tests (extracted 13^th^ May 2020)

https://www.worldometers.info/coronavirus/

## Drugs

We attempted to get data for lipid-lowering drug consumption, but as these data were sparse and not systematically comparable, we desisted from further attempts.

## Mercury

Country wide mercury consumption is not easily available. UN-EN reports give tons of consumption for those countries that use mercury in the chlorine-alkaline industry but this is only about half of the countries. For the rest only regional data were available. We contacted Dr. Steenhuisen and Prof. Kümmerer in the hope to get help. Dr. Steenhuisen did not provide data and the publication (Steenhuisen, F., & Wilson, S. J. (2019). Development and application of an updated geospatial distribution model for gridding 2015 global mercury emissions. *Atmospheric Environment, 211*, 138-150.

doi:https://doi.org/10.1016/j.atmosenv.2019.05.003) did not contain data. Prof. Kümmerer, an expert in environmental toxicology did not have any information, neither did other experts asked. Hence we dropped this variable in the final analysis, as the data from the UN-EN report on mercury in the alkaline industry was too patchy.

At each step new partial data-sets were created and added to the master database with the Statistica-data “merge” command with variable merging according to country names and each set was saved under a new name.

Since some variables were created as functions of others and indexed by the original variable number, the final database contained all variables, even those that were never further used except for orienting correlations.

## Transformations

Outcome variables were log-transformed to check whether they would then be normally distributed to allow for a normal linear regression. As the result was less than satisfactory we decided to go for a generalized model and regress on a gamma-distributed variable, as the variables were clearly gamma-distributed.

Date variables were calculated from starting date to the 15^th^ May for border closure, school closure, and finally for the rapidity of reaction as the difference. This was defined as the date when the first measure, either border closure or school closure was registered and the days to first case registration was calculated.

## First Analyses

Distribution analyses of the dependent variable showed that it was gamma distributed and that a log transformation cannot rectify this. Hence it was decided that a regression on gamma-distributed variables should be calculated.

First non-parametric correlations were correlated to see which variables correlate at all with the outcome, and as defined in the protocol, only variables with r > .3 and/or significantly correlating variables were considered further in regression models. The regression models used the functionality Generalized Linear Models, stipulating a gamma distributed outcome variable with a log-link function. The parametrization method was overparametrized, as there was no sigma restricted coding in our data.

All potentially included variables were inspected for their descriptive parameters and to see, whether they contribute any variance. If several similar variables (.e.g. air pollution variables) were available we used those that had the least missing data in order to not lose power.

All modeling approaches included first health serviced, population and health parameters in a model, trying to fit the most parsimonious model with only significant predictors in the equation. This was done by calculating forced entry models, excluding non-significant predictors and recalculating the model. Highly intercorrelated variables were never used together, but separate models were calculated and the model with the best model fit was selected.

After that variables representing political actions (country lockdown, school closure) were entered in an additional model and used if significant as predictors.

## References

1. Pan A, Liu L, Wang C, Guo H, Hao X, Wang Q, et al. Association of Public Health Interventions With the Epidemiology of the COVID-19 Outbreak in Wuhan, China. JAMA. 2020;online first. doi: 10.1001/jama.2020.6130.

2. Tang X, Wu C, Li X, Song Y, Yao X, Wu X, et al. On the origin and continuing evolution of SARS-CoV-2. National Science Review. 2020. doi: 10.1093/nsr/nwaa036.

3. Wang X, Xu W, Hu G, Xia S, Sun Z, Liu Z, et al. SARS-CoV-2 infects T lymphocytes through its spike protein-mediated membrane fusion. Cellular & Molecular Immunology. 2020. doi: 10.1038/s41423-020-0424-9.

4. Wang K, Chen W, Zhou Y-S, Lian J-Q, Zhang Z, Du P, et al. SARS-CoV-2 invades host cells via a novel route: CD147-spike protein. bioRxiv. 2020:2020.03.14.988345. doi: 10.1101/2020.03.14.988345.

5. Zheng M, Gao Y, Wang G, Song G, Liu S, Sun D, et al. Functional exhaustion of antiviral lymphocytes in COVID-19 patients. Cellular & Molecular Immunology. 2020. doi: 10.1038/s41423-020-0402-2.

6. Grasselli G, Zangrillo A, Zanella A, Antonelli M, Cabrini L, Castelli A, et al. Baseline Characteristics and Outcomes of 1591 Patients Infected With SARS-CoV-2 Admitted to ICUs of the Lombardy Region, Italy. JAMA. 2020;323(16):1574–81. doi: 10.1001/jama.2020.5394.

7. Bi Q, Wu Y, Mei S, Ye C, Zou X, Zhang Z, et al. Epidemiology and transmission of COVID-19 in 391 cases and 1286 of their close contacts in Shenzhen, China: a retrospective cohort study. The Lancet Infectious Diseases. 2020. doi: 10.1016/S1473-3099(20)30287-5.

8. Dehning J, Zierenberg J, Spitzner FP, Wibral M, Neto JP, Wilczek M, et al. Inferring change points in the spread of COVID-19 reveals the effectiveness of interventions. Science. 2020:eabb9789. doi: 10.1126/science.abb9789.

9. Ben-Israel I. The end of exponential growth: The decline in the spread of the coronavirus Jerusalem: Times of Israel; 2020 [cited 2020 22nd April]. Available from: https://www.timesofisrael.com/the-end-of-exponential-growth-the-decline-in-the-spread-of-coronavirus/.

10. Kuhbandner C. Von der fehlenden wissenschaftlichen Begründung der Corona-Maβnahmen Heidelberg: Spektrum; 2020 [cited 2020 27.4.]. Available from: https://scilogs.spektrum.de/menschen-bilder/von-der-fehlenden-wissenschaftlichen-begruendung-der-corona-massnahmen/.

11. Ferguson N, Laydon D, Nedjati Gilani G, Imai N, Ainslie K, Baguelin M, et al. Impact of non-pharmaceutical interventions (NPIs) to reduce COVID19 mortality and healthcare demand. London: Imperial College, 2020.

12. An der Heiden M, Buchholz U. Modellierung von Beispielszenarien der SARS-CoV-2-Epidemie 2020 in Deutschland. Berlin: Robert Koch Institut, 2020.

13. Gomes MGM, Corder RM, King JG, Langwig KE, Souto-Maior C, Carneiro J, et al. Individual variation in susceptibility or exposure to SARS-CoV-2 lowers the herd immunity threshold. medRxiv. 2020:2020.04.27.20081893. doi: 10.1101/2020.04.27.20081893.

14. Lewis N. Why herd immunity to COVID-19 is reached much earlier than thought 2020. Available from: https://judithcurry.com/2020/05/10/why-herd-immunity-to-covid-19-is-reached-much-earlier-than-thought/.

15. Burnham KP, Anderson DR. Model Selection and Inference: A Practical Information-Theoretic Approach. York N, editor 1998.

16. McQuarrie ADR, Tsai C-L. Regression and time-series model selection. Singapore: World Scientific Publishers; 1998.

17. Schlage WK, Westra JW, Gebel S, Catlett NL, Mathis C, Frushour BP, et al. A computable cellular stress network model for non-diseased pulmonary and cardiovascular tissue. BMC Systems Biology. 2011;5(1):1–15. doi: 10.1186/1752-0509-5-168.

18. Wiegant FAC, Souren JEM, van Wijk R. Stimulation of survival capacity in heat shocked cells by subsequent exposure to minute amounts of chemical stressors; role of similartiy in hsp-inducing effects. Human and Experimental Toxicology. 1999;18:460–70.

19. Israel A, Feldhamer I, Lahad A, Levin-Zamir D, Lavie G. Smoking and the risk of COVID-19 in a large observational population study. medRxiv. 2020:2020.06.01.20118877. doi: 10.1101/2020.06.01.20118877.

20. Hopkinson NS, Rossi N, El-Sayed Moustafa J, Laverty AA, Quint JK, Freydin MB, et al. Current tobacco smoking and risk from COVID-19: results from a population symptom app in over 2.4 million people. medRxiv. 2020:2020.05.18.20105288. doi: 10.1101/2020.05.18.20105288.

21. An der Heiden M, Hamouda O. Schätzung der aktuellen Entwicklung der SARS-CoV-2-Epidemie in Deutschland – Nowcasting. Epidemiologisches Bulletin. 2020;17:10–5. doi: 10.25646/6692.2.

22. Tolksdorf E, Buda S, Schuler E, Wieler LH, Haas W. Schwereeinschätzung von COVID-19 mit Vergleichsdaten zu Pneumonien aus dem Krankenhaussentinel für schwere akute Atemwegserkrankungen am RKI (ICOSARI). Epidemiologisches Bulletin. 2020;14:3–9. doi: 10.25646/6601.

23. Moran RJ, Fagerholm ED, Daunizeau J, Cullen M, Richardson MP, Williams S, et al. Estimating required lockdown cycles before immunity to SARS-CoV-2: Model-based analyses of susceptible population sizes, S0, in seven European countries including the UK and Ireland. medRxiv. 2020:2020.04.10.20060426. doi: 10.1101/2020.04.10.20060426.

24. Friston KJ, Parr T, Zeidman P, Razi A, Flandin G, Daunizeau J, et al. Tracking and tracing in the UK: a dynamic causal modeling study. arXiv. 2020;2005.07994.

25. Patel JJ. The things we say. JAMA. 2018;319(4):341–2. doi: 10.1001/jama.2017.20545.

26. Colloca L, Barsky AJ. Placebo and nocebo effects. New England Journal of Medicine. 2020;382(6):554–61.

27. Flaxman S, Mishra S, Gandy A, Unwin HJT, Mellan TA, Coupland H, et al. Estimating the effects of non-pharmaceutical interventions on COVID-19 in Europe. Nature. 2020. doi: 10.1038/s41586-020-2405-7.

28. Brauner JM, Mindermann S, Sharma M, Stephenson AB, Gavenčiak T, Johnston D, et al. The effectiveness and perceived burden of nonpharmaceutical interventions against COVID-19 transmission: a modeling study with 41 countries. medRxiv. 2020:2020.05.28.20116129. doi: 10.1101/2020.05.28.20116129.

29. Ioannidis J. The infection fatality rate of COVID-19 inferred from seroprevalence data. medRxiv. 2020:2020.05.13.20101253. doi: 10.1101/2020.05.13.20101253.

30. Daunizeau J, Moran RJ, Mattout J, Friston K. On the reliability of model-based predictions in the context of the current COVID epidemic event: impact of outbreak peak phase and data paucity. medRxiv. 2020:2020.04.24.20078485. doi: 10.1101/2020.04.24.20078485.

31. Chinazzi M, Davis JT, Ajelli M, Gioannini C, Litvinova M, Merler S, et al. The effect of travel restrictions on the spread of the 2019 novel coronavirus (COVID-19) outbreak. Science. 2020:eaba9757. doi: 10.1126/science.aba9757.

32. Tian H, Liu Y, Li Y, Wu C-H, Chen B, Kraemer MUG, et al. An investigation of transmission control measures during the first 50 days of the COVID-19 epidemic in China. Science. 2020:eabb6105. doi: 10.1126/science.abb6105.

33. Valenti L, Bergna A, Pelusi S, Facciotti F, Lai A, Tarkowski M, et al. SARS-CoV-2 seroprevalence trends in healthy blood donors during the COVID-19 Milan outbreak. medRxiv. 2020:2020.05.11.20098442. doi: 10.1101/2020.05.11.20098442.

34. Cheng H-Y, Jian S-W, Liu D-P, Ng T-C, Huang W-T, Lin H-H, et al. Contact Tracing Assessment of COVID-19 Transmission Dynamics in Taiwan and Risk at Different Exposure Periods Before and After Symptom Onset. JAMA Internal Medicine. 2020. doi: 10.1001/jamainternmed.2020.2020.

35. Cowling BJ, Ali ST, Ng TWY, Tsang TK, Li JCM, Fong MW, et al. Impact assessment of non-pharmaceutical interventions against coronavirus disease 2019 and influenza in Hong Kong: an observational study. The Lancet Public Health. 2020. doi: 10.1016/S2468-2667(20)30090-6.

36. Hellewell J, Abbott S, Gimma A, Bosse NI, Jarvis CI, Russell TW, et al. Feasibility of controlling COVID-19 outbreaks by isolation of cases and contacts. The Lancet Global Health. 2020;8(4):e488–e96. doi: 10.1016/S2214-109X(20)30074-7.

37. Kucharski AJ, Russell TW, Diamond C, Liu Y, Edmunds J, Funk S, et al. Early dynamics of transmission and control of COVID-19: a mathematical modeling study. The Lancet Infectious Diseases. 2020;20(5):553–8. doi: 10.1016/S1473-3099(20)30144-4.

38. Walach H, Hockertz S. Wuhan Covid19 data – more questions than answers. Toxicology. 2020;440:152486. doi: https://doi.org/10.1016/j.tox.2020.152486.

39. Kuhbandner C, Homburg S, Walach H, Hockertz S. Comment on Dehning et al (Science, 15 May 2020, eabb9789: Inferring change points in the spread of COVID-19 reveals the effectiveness of interventions). advance Social Sciences and Humanities Preprint. 2020;Preprint. doi: https://doi.org/10.31124/advance.12362645.v1.

40. Hains DS, Schwaderer AL, Carroll AE, Starr MC, Wilson AC, Amanat F, et al. Asymptomatic Seroconversion of Immunoglobulins to SARS-CoV-2 in a Pediatric Dialysis Unit. JAMA. 2020. doi: 10.1001/jama.2020.8438.

41. Edridge AW, Kaczorowska JM, Hoste AC, Bakker M, Klein M, Jebbink MF, et al. Human coronavirus reinfection dynamics: lessons for SARS-CoV-2. medRxiv. 2020:2020.05.11.20086439. doi: 10.1101/2020.05.11.20086439.

42. Schäfer A, Baric RS. Epigenetic landscape during coronavirus infection. Pathogens. 2017;6(1):8. PubMed PMID: doi:10.3390/pathogens6010008.

43. Totura AL, Baric RS. SARS coronavirus pathogenesis: host innate immune responses and viral antagonism of interferon. Current Opinion in Virology. 2012;2(3):264-75. doi: https://doi.org/10.1016/j.coviro.2012.04.004.

44. Shayakhmetov DM, Di Paolo NC, Mossman KL. Recognition of virus infection and innate host responses to viral gene therapy vectors. Molecular Therapy. 2010;18(8):1422–9. doi: 10.1038/mt.2010.124.

45. Friston KJ, Parr T, Zeidman P, Razi A, Flandin G, Daunizeau J, et al. Second waves, social distancing, and the spread of COVID-19 across America. arxiv. 2020;1104.3344v1.

46. Breen R, Karlson KB, Holm A. Interpreting and understanding logits, probits, and other nonlinear probability models. Annual Review of Sociology. 2018;44:4.1-.16. doi: 10.1146/annurev-soc-073117-04142.

47. Richards SA, Whittingham MJ, Stephens PA. Model selection and model averaging in behavioural ecology: the utility of the IT-AIC framework. Behavioural Ecology and Sociobiology. 2011;65:77–89. doi: 10.1007/s00265-010-1035-8.

48. Kuha J. AIC and BIC: Comparisons of assumptions and performance. Sociological Methods and Research. 2004;33:188–229.

49. Wagenmakers EJ, Farrell S. AIC model selection using Akaike weights. Psychonomic Bulletin & Review. 2004;11:192–6.

50. Claeskens G, Hjort NL. Model Selection and Model Averaging. Cambridge: Cambridge University Press; 2008.

## Other Sources on Health and Population Data

EMB-Japan. (2012). Japan und Deutschland im Zahlenvergleich (2): Bevölkerung. Accessed 15. May 2020 from https://www.de.emb-japan.go.jp/NaJ/NaJ1202/dj2.html

Hurriyet. (2020). Türkei: Mehr Einpersonenhaushalte und kleinere Haushaltsgröβen. Accessed 15. May 2020 from https://www.hurriyet.de/news_tuerkei-mehr-einpersonenhaushalte-und-kleinere-haushaltsgroessen92527_143536701.html

International Diabetes Federation. (2020). IDF Europe members. Accessed 15. May 2020 fromInternational Diabetes Federation: https://idf.org/our-network/regions-members/europe/members.html

International Diabetes Federation. (2020). IDF MENA Members. Accessed 15. May 2020 fromInternational Diabetes Federation: https://idf.org/our-network/regions-members/middle-east-and-north-africa/members.html

International Diabetes Federation. (2020). IDF North America and Caribbean members. Accessed 15. May 2020 from International Diabetes Federation: https://idf.org/our-network/regions-members/north-america-and-caribbean/members.html

International Diabetes Federation. (2020). IDF SACA members. Accessed 15. May 2020 fromInternational Diabetes Federation: https://idf.org/our-network/regions-members/south-and-central-america/members.html

International Diabetes Federation. (2020). IDF SEA members. Accessed 15. May 2020 fromInternational Diabetes Federation: https://idf.org/our-network/regions-members/south-east-asia/members.html

International Diabetes Federation. (2020). IDF Western Pacific members. Accessed 15. May 2020 fromInternational Diabetes Federation: https://idf.org/our-network/regions-members/western-pacific/members.html

Laenderdaten. (2020). Bevölkerungsdichte nach Ländern. Accessed 15. May 2020 from https://www.laenderdaten.info/bevoelkerungsdichte.php

Our World in Data. (2020). Do more people live in urban or rural areas? Accessed 15. May 2020 fromOur World in Data: https://ourworldindata.org/urbanization#all-charts-preview

Statista. (2018). Anteil der Einpersonenhaushalte an allen Privathaushalten in den Ländern der EU im Jahr 2017. Accessed 15. May 2020 fromStatista: https://de.statista.com/statistik/daten/studie/156448/umfrage/anteil-der-einpersonenhaushalte-an-allen-privathaushalten-in-den-laendern-der-eu-im-jahr-2009/

Statista. (2019). China: Altersstruktur von 2008 bis 2018. Accessed 15. May 2020 fromStatista: https://de.statista.com/statistik/daten/studie/166164/umfrage/altersstruktur-in-china/

Statista. (2019). Groβbritannien: Altersstruktur von 2008 bis 2018. Accessed 15. May 2020 from Statista: https://de.statista.com/statistik/daten/studie/167292/umfrage/altersstruktur-in-grossbritannien/

Statista. (2019). Japan: Altersstruktur von 2008 bis 2018. Accessed 15. May 2020 fromStatista: https://de.statista.com/statistik/daten/studie/165976/umfrage/altersstruktur-in-japan/

Statista. (2019). Norwegen: Altersstruktur von 2008 bis 2018. Accessed 15. May 2020 fromStatista: https://de.statista.com/statistik/daten/studie/258694/umfrage/altersstruktur-in-norwegen/

Statista. (2019). Prävalenz von Diabetes bei zwischen 20-und 79-Jährigen in ausgewählten Ländern weltweit im Jahr 2019. Accessed 15. May 2020 fromStatista: https://de.statista.com/statistik/daten/studie/182587/umfrage/praevalenz-von-diabetes-in-ausgewaehlten-laendern/

Statista. (2019). Russland: Altersstruktur von 2008 bis 2018. Accessed 15. May 2020 fromStatista: https://de.statista.com/statistik/daten/studie/171397/umfrage/altersstruktur-in-russland/

Statista. (2019). Türkei: Altersstruktur von 2008 bis 2018. Accessed 15. May 2020 fromStatista: https://de.statista.com/statistik/daten/studie/216075/umfrage/altersstruktur-in-der-tuerkei/

Statista. (2019). USA: Altersstruktur von 2008 bis 2018. Accessed 15. May 2020 fromStatista: https://de.statista.com/statistik/daten/studie/165801/umfrage/altersstruktur-der-usa/

Statista. (April 2020). Altersstruktur der ständigen Wohnbevölkerung in der Schweiz von 2009 bis 2019. Accessed 15. May 2020 fromStatista: https://de.statista.com/statistik/daten/studie/216731/umfrage/altersstruktur-in-der-schweiz/

Statista. (2020). Brasilien: Altersstruktur von 2008 bis 2018. Accessed 15. May 2020 fromStatista: https://de.statista.com/statistik/daten/studie/169267/umfrage/altersstruktur-in-brasilien/

Statista. (März 2020). Europäische Union: Altersstruktur in den Mitgliedsstaaten im Jahr 2019. Accessed 15. May 2020 fromStatista: https://de.statista.com/statistik/daten/studie/248981/umfrage/altersstruktur-in-den-eu-laendern/

Statista. (May 2020). Europäische Union: Bevölkerungsdichte in den Mitgliedsstaaten im Jahr 2018. Accessed 15. May 2020 fromStatista: https://de.statista.com/statistik/daten/studie/74693/umfrage/bevoelkerungsdichte-in-den-laendern-der-eu/

Statista. (2020). Indien: Altersstruktur von 2008 bis 2018. Accessed 15. May 2020 fromStatista: https://de.statista.com/statistik/daten/studie/170740/umfrage/altersstruktur-in-indien/

Statista. (2020). Iran: Altersstruktur von 2008 bis 2018. Accessed 15. May 2020 fromStatista: https://de.statista.com/statistik/daten/studie/259400/umfrage/altersstruktur-im-iran/

Statista. (2020). Kanada: Altersstruktur von 2008 bis 2018. Accessed 15. May 2020 fromStatista: https://de.statista.com/statistik/daten/studie/77192/umfrage/altersstruktur-in-kanada/

Wikipedia. (2020). Liste der Länder nach Geschlechterverteilung. Accessed 15. May 2020 from https://de.wikipedia.org/wiki/Liste_der_L%C3%A4nder_nach_Geschlechterverteilung

Zeit. (2013). Schweizer, allein, glücklich: Allein zu Haus. Accessed 15. May 2020 from https://www.zeit.de/2013/21/alleine-wohnen-schweiz

